# Pharmacovigilance organization and training needs of health personnels in health facilities of Cameroon: a cross-sectional study

**DOI:** 10.64898/2026.08.26.26361382

**Authors:** Augustin Murhabazi Bashombwa, Ketina Hirma Tchio-Nighie, Merveille Claire Nana Djapou, Collins BUH Nkum, Iyakachi Blama Abba, Cavin Epie Bekolo, Jerome Ateudjieu

## Abstract

Health facilities (HFs) routinely administer medicines and are expected to ensure patient safety by detecting, reporting, investigating, and analysing adverse events following exposure to drugs (AEFED). This study aimed to assess the implementation of pharmacovigilance activities in referral and regional health facilities in Cameroon and to identify pharmacovigilance training needs among healthcare personnel (HP). This was a cross-sectional descriptive study targeting referral and regional health facilities and healthcare personnel involved in patient care and pharmacovigilance activities in Cameroon. Health facilities were selected using stratified purposive sampling, while healthcare personnel were selected through exhaustive sampling. Data were collected using semi-structured electronic questionnaires administered face-to-face by trained enumerators. The questionnaires assessed the organization, resources, and implementation of pharmacovigilance activities at health facilities, as well as healthcare personnel knowledge of pharmacovigilance concepts, previous training, and perceived training needs. Of the 14 eligible health facilities, 10 (71.4%) consented to participate in the study. Of the 10 health facilities, 4 (40.0%) had an established pharmacovigilance unit, while 3 (30.0%) reported conducting neither detection nor notification activities. Among the 261 healthcare personnel approached, 214 (81.9%) participated. Only 41.6% had needed knowledge to detect an adverse event, while 72.9% were aware of adverse event notification procedures. Previous exposure to pharmacovigilance training was reported by 37.9% of healthcare personnel, and all participants expressed a need for additional training, particularly on national pharmacovigilance regulations (69.2%), organization of the pharmacovigilance system (67.3%), and adverse event detection (67.3%). The main reported challenges by healthcare personnel in the implementation of pharmacovigilance activities included insufficient budget allocation, limited access to pharmacovigilance training, lack of pharmacovigilance guidelines and insufficient qualified human resources. Pharmacovigilance implementation in referral and regional health facilities in Cameroon remains limited, with gaps in organizational structures, resources, healthcare personnel knowledge, and training. Strengthening pharmacovigilance systems through improved facility capacity, availability of essential tools, and targeted healthcare personnel training is needed to enhance drug safety surveillance.

## Background

Most of marketed therapeutic products include from clinical trials needed information to assess it benefit-risk ratio [1]. However, it use in routine care significantly increases the number of people exposed to the product, thereby increasing the likelihood of detecting rare adverse events or longer-term effects of the product, which may require a reassessment of the benefit-risk ratio and the resulting decisions [2, 3]. Similarly, manufacturing defects, failures to maintain proper storage conditions throughout the supply chain, using the products under conditions different from those of clinical trials, and the use of counterfeit medicines can all generate adverse events that must be detected and prevented [4].

Health systems, health program, and hospitals administering or using therapeutic products are recommended to establish a pharmacovigilance system to detect, report, investigate, and conduct causality assessments, in order to generate the evidence needed for decision-making aimed at minimizing the risks of population exposure to these products [5-7].

According to the World Health Organization (WHO), approximately 1 in 30 patients experiences adverse drug reactions during treatment, and more than a quarter of these reactions are considered serious or life-threatening [8]. Furthermore, a worldwide conducted systematic review found that approximately 17% of patients experience a medication-related adverse event during their hospitalization [9]. In Africa, the burden is equally significant, with a systematic review reporting that approximately 7.8% of hospitalized patients experienced a drug-related adverse event [10].

WHO has developed and made accessible guidelines for implementation of pharmacovigilance in health facilities [11]. In line with the guideline, Cameroon ministry of public health has developed guidelines organizing drug safety monitoring in health progams [12, 13]. However, despite the availability of the guideline the performances of the pharmacovigilance are still under expectations.

## Materials and Methods

### Ethical considerations

Study objectives, procedures, and requirements were explained to the heads of participating health facilities and to targeted healthcare personnel prior to data collection. Written informed consent was obtained from all participants before questionnaire administration, and only consenting individuals were enrolled. The study protocol was reviewed and approved by the Cameroon National Ethics Committee for Human Health Research (Approval No. N°2020/10/1305/CE/CNERSH/SP).

### Study design

This was a cross-sectional descriptive study targeting referral and regional health facilities of Cameroon selected by purposive sampling and healthcare personnel selected by exhaustive sampling in included health facilities. Data were collected by trained enumerators using an electronic semi-structured questionnaire administered face-to-face to key healthcare personnel responsible for medicine safety monitoring or drug management, as well as to healthcare personnel to assess pharmacovigilance knowledge and training needs.

### Study period and setting

The study was conducted from 1^st^ November to 31^th^ December 2020 in referral and regional health facilities in the Centre, West, Adamawa, and Littoral regions of Cameroon. Referral health facilities were defined as hospitals providing the highest level of specialized care at the national level, whereas regional health facilities represented the highest level of referral care within each country’s region.

### Study population and sampling

Eligible health facilities were those classified as referral or regional-level facilities within the Cameroonian health system, as listed in the Medical Directory of the Ministry of Public Health of Cameroon [14]. We included Referral and regional health facilities that authorized study implementation. Non-functional health facilities were excluded.

Eligible healthcare personnel included pharmacovigilance focal points, physicians, nurses, other healthcare providers involved in outpatient care services, and pharmacists engaged in the supply or management of medical products for patients. Healthcare personnel who provided informed consent were included. Interns and personnel on leave for more than six months were excluded.

### Data collection tools and variables

Data collection tools were developed based on the study objectives and were pretested for validation before data collection. For health facilities, key variables assessed included the existence of pharmacovigilance units, availability of infrastructure and essential resources, implementation of core pharmacovigilance activities, and perceived needs for strengthening pharmacovigilance. For healthcare personnel, data were collected on pharmacovigilance knowledge, previous exposure to pharmacovigilance training, training needs, and perceived challenges related to the implementation of pharmacovigilance activities.

### Data collection process

In each facility, the head was briefed on study objectives, procedures, and eligibility criteria, then identified appropriate respondents. Trained surveyors obtained informed consent before administering the facility questionnaire. For the healthcare personnel questionnaire, eligible individuals were informed about the study and, after providing written consent, completed it face-to-face with trained surveyors.

### Data management

Field supervisors reviewed data daily for completeness and consistency before database upload. Qualified data managers subsequently downloaded, cleaned, and corrected the data, resolving errors and inconsistencies in collaboration with field teams.

### Data analysis

Data were analysed using Stata 17 (StataCorp LLC, College Station, TX, USA). Descriptive analyses (frequencies, proportions) assessed health facilities’ pharmacovigilance units, infrastructure, equipment, tools, core activities (detection, notification, data analysis), and perceived needs. Among healthcare personnel, analyses assessed pharmacovigilance knowledge, prior training exposure, and perceived training needs.

## Results

### 1. Characteristics of health facilities and healthcare personnels

Of the 14 eligible health facilities, 10 (71.4%) agreed to participate, including four referral hospitals and six regional hospitals. Among the 261 healthcare personnel approached, 214 (81.9%) consented to participate.

Table 1 summarizes the characteristics of the participating healthcare personnel. Nurses constituted the largest professional group (44.9%), followed by physicians (29.0%) and pharmacists (21.5%). More than half of the participants were female (56.1%), and 41.1% had more than 10 years of professional experience.

**Table 1:**
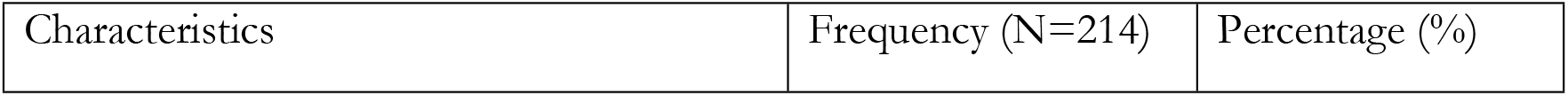
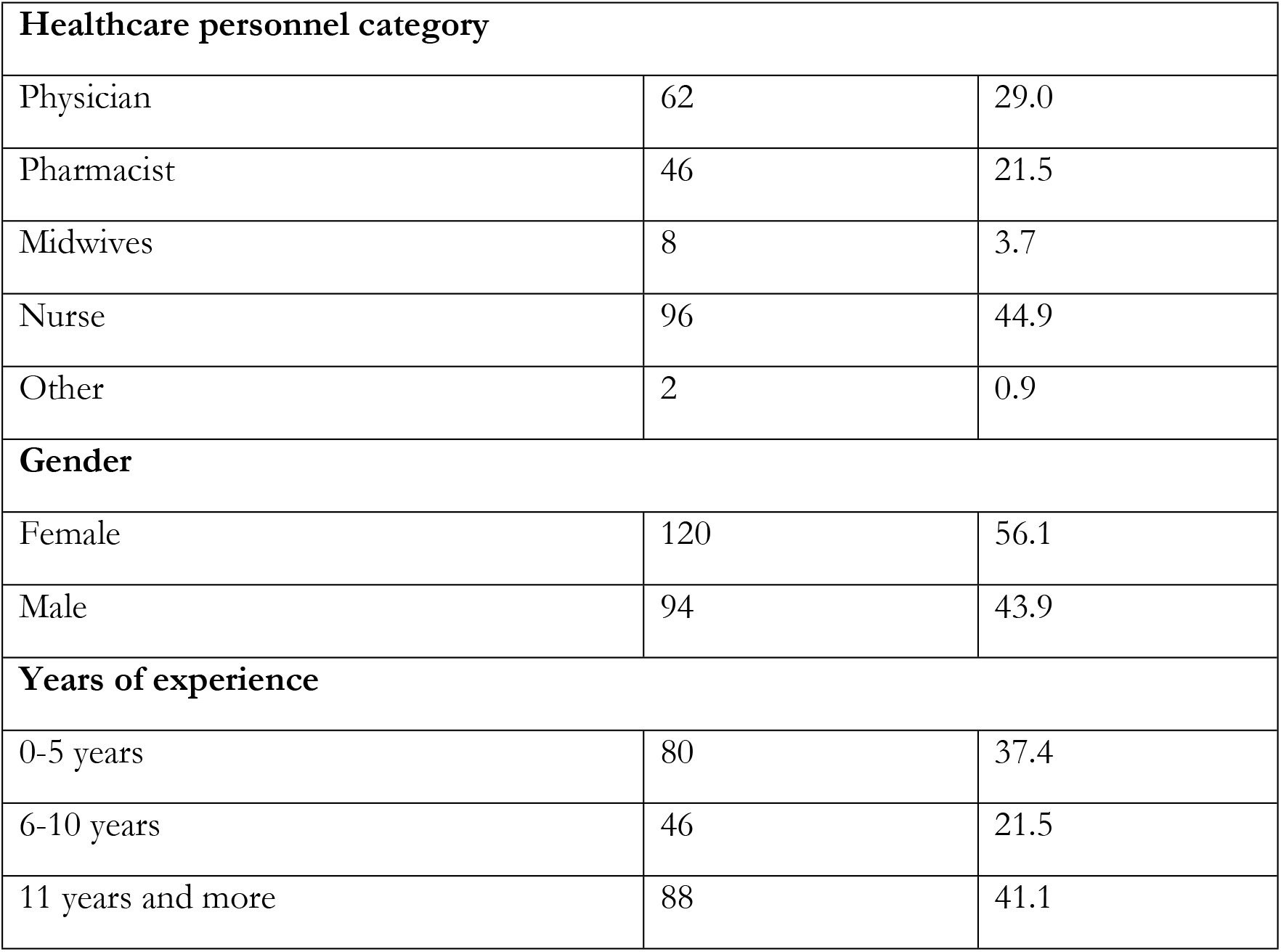
Distribution of healthcare personnel per category and type of health facilities.

| Characteristics | Frequency (N=214) | Percentage (%) |
| --- | --- | --- |

| **Healthcare personnel category** |  |  |
| --- | --- | --- |
| Physician | 62 | 29.0 |
| Pharmacist | 46 | 21.5 |
| Midwives | 8 | 3.7 |
| Nurse | 96 | 44.9 |
| Other | 2 | 0.9 |
| **Gender** |  |  |
| Female | 120 | 56.1 |
| Male | 94 | 43.9 |
| **Years of experience** |  |  |
| 0-5 years | 80 | 37.4 |
| 6-10 years | 46 | 21.5 |
| 11 years and more | 88 | 41.1 |

### 2. Distribution of pharmacovigilance activities in health facilities

Table 2 presents the distribution of pharmacovigilance activities in health facilities. Of the 10 health facilities, 4 (40.0%) were reported to have an established pharmacovigilance unit, while 3 (30.0%) reported conducting neither detection nor notification activities. Only 3 (30.0%) health facilities reported to conduct analysis of pharmacovigilance data.

**Table 2:** Distribution of pharmacovigilance activities in health facilities.

| **Variables** | **Frequency** | **Proportion (%)** |
| --- | --- | --- |
| **Pharmacovigilance Unit** |  |  |
| Yes | 4 | 40.0 |
| No | 6 | 60.0 |
| **Pharmacovigilance activities** |  |  |
| Yes | 7 | 70.0 |
| No | 3 | 30.0 |
| **Structure to which notification is done** |  |  |
| Health district | 2 | 20.0 |
| Regional Delegation of Public Health | 4 | 40.0 |
| Directorate of Pharmacy, Medicines and Laboratories (DPML) | 3 | 30.0 |
| We do not transfer data | 3 | 30.0 |
| **Notification delay** |  |  |
| Less than one week | 3 | 30.0 |
| Less than one month | 3 | 30.0 |
| Less than three months | 1 | 10.0 |
| Not applicable (no data transmission) | 3 | 30.0 |
| **Frequency of pharmacovigilance data analysis** |  |  |
| None | 7 | 70.0 |
| Monthly | 1 | 10.0 |
| Quarterly | 1 | 10.0 |
| Semestral | 1 | 10.0 |
| **Analyses performed on pharmacovigilance data** |  |  |
| Completeness of reporting sites | 1 | 10.0 |
| Completeness of notification forms | 2 | 20.0 |
| Timeliness of notification form submission | 3 | 30.0 |
| Reporting rate | 1 | 10.0 |
| Investigation rate | 1 | 10.0 |
| Frequency analysis | 1 | 10.0 |
| Causality assessment | 1 | 10.0 |
| Reporting quality assessment | 1 | 10.0 |
| List of medicines to be circulated in the hospital, medicine orders, and expiry dates | 1 | 10.0 |
| None | 7 | 70.0 |
| **Feedback on data analysis** |  |  |
| No | 8 | 80.0 |
| Yes | 2 | 20.0 |
| **Recipients of feedback on data analysis** |  |  |
| Reporting service/unit | 1 | 50.0 |
| Health district | 1 | 50.0 |
| Directorate of Pharmacy, Medicines and Laboratories (DPML) | 1 | 50.0 |

### 3. Availability of tools and resources for pharmacovigilance in health facilities

Table 3 present the available tools and resources for pharmacovigilance in health facilities. Half of the facilities (50.0%) reported having no pharmacovigilance resources, while trained personnel were available in five facilities (50.0%). Data analysis software, data archiving systems, computers, and telephone lines were each available in only one facility (10.0%). Notification forms were the most commonly available pharmacovigilance tool (70.0%), whereas pharmacovigilance guidelines, investigation forms, and detection forms were each available in only one facility (10.0%).

### 4. Previous exposure of healthcare personnels to training in pharmacovigilance

Table 4 presents previous exposure of healthcare personnels to training in pharmacovigilance. Overall, 81 of the 214 (37.9%) healthcare personnel reported having previously received pharmacovigilance training. The training most frequently covered adverse event detection (50.6%), adverse event reporting (46.7%), and Good Clinical Practice (GCP) in pharmacovigilance (32.1%).

**Table 3:** Availability of resources, and tools for pharmacovigilance in health facilities.

| Variables | Frequency | Proportion (%) |
| --- | --- | --- |
| **Health facility resources for pharmacovigilance** |  |  |
| Trained personnel | 5 | 50.0 |
| Data analysis software | 1 | 10.0 |
| Data archiving system | 1 | 10.0 |
| Computer | 1 | 10.0 |
| Telephone line | 1 | 10.0 |
| None | 5 | 50.0 |
| **Pharmacovigilance tools available at the health facility** |  |  |
| Pharmacovigilance guideline | 1 | 10.0 |
| Notification form | 7 | 70.0 |
| Investigation form | 1 | 10.0 |
| Detection form | 1 | 10.0 |
| None | 3 | 30.0 |

**Table 4:** Previous exposure of health personnels to pharmacovigilance training.

| Variables | Frequency (N=214) | Percentage (%) |
| --- | --- | --- |
| **Received previous pharmacovigilance training** | 81 | 37.9 |
| **Source of previous training** |  |  |
| Online course (e-learning) | 4 | 4.9 |
| Workshops | 25 | 30.9 |
| Internship | 8 | 9.9 |
| Degree courses | 48 | 59.3 |
| Other | 12 | 14.8 |
| **Training topics covered** |  |  |
| National pharmacovigilance regulations | 19 | 23.5 |
| International pharmacovigilance regulations | 21 | 25.9 |
| Organization of the pharmacovigilance system in Cameroon | 13 | 16.0 |
| Adverse event detection | 41 | 50.6 |
| Adverse event reporting | 37 | 46.7 |
| Data analysis | 13 | 16.0 |
| Causality assessment | 9 | 11.1 |
| Good Clinical Practice (GCP) in Pharmacovigilance | 26 | 32.1 |
| Other | 19 | 23.5 |

### 5. Knowledge of healthcare personnels on pharmacovigilance

Table 5 presents healthcare personnel’s knowledge of pharmacovigilance. Overall, 89 (41.6%) participants correctly defined an adverse event, including 46.8% of physicians, 43.5% of pharmacists, 34.4% of nurses, and 62.5% of midwives. Regarding reporting procedures, 156 (72.9%) participants knew how to report adverse events, while 151 (70.6%) knew to whom adverse events should be reported.

**Table 5:** Knowledge of healthcare personnel on pharmacovigilance.

| Variables | Nurse<br>(N=96) | Physician<br>(N=62) | Pharmacist<br>(N=46) | Midwives<br>(N=8) | Others<br>(N=2) | Overall<br>(N=214) |
| --- | --- | --- | --- | --- | --- | --- |
| Knows the definition of an adverse event | 33 (34.4) | 29 (46.8) | 20 (43.5) | 5 (62.5) | 2 (100.0) | 89 (41.6) |
| Know the definition of a serious adverse event | 25 (26) | 20 (32.3) | 14 (30.4) | 4 (50) | 2 (100.0) | 65 (30.4) |
| Know the definition of a minor adverse event | 85 (88.5) | 56 (90.3) | 35 (76.1) | 7 (87.5) | 2 (100.0) | 185 (86.5) |
| Know why notification of adverse events is done | 87 (90.6) | 59 (95.2) | 45 (97.8) | 7 (87.5) | 2 (100.0) | 200 (93.5) |
| Know reporting procedures | 75 (78.1) | 45 (72.6) | 29 (63) | 5 (62.5) | 2 (100.0) | 156 (72.9) |
| Knows the notification delay | 86 (89.6) | 53 (85.5) | 38 (82.6) | 8 (100) | 2 (100.0) | 187 (87.4) |
| Knows who is responsible for notification | 68 (70.8) | 49 (79) | 37 (80.4) | 7 (87.5) | 1 (50.0) | 162 (75.7) |
| Knows to whom adverse events should be notified | 56 (58.3) | 51 (82.3) | 38 (82.6) | 5 (62.5) | 1 (50.0) | 151 (70.6) |

### 6. Pharmacovigilance training needs of health personnels and challenges to implementation

Table 6 presents the pharmacovigilance training needs and the challenges encountered during the implementation of pharmacovigilance activities. All healthcare personnel reported a need for training in pharmacovigilance. The most frequently requested training topics were the national regulations governing pharmacovigilance in Cameroon (69.2%) and adverse event detection (67.3%). The most commonly reported barrier to the implementation of pharmacovigilance activities was the lack of pharmacovigilance guidelines (58.9%).

**Table 6:** Pharmacovigilance training needs of healthcare personnel and challenges to pharmacovigilance implementation.

| Variables | Frequency<br>(N=214) | Percentage<br>(%) |
| --- | --- | --- |
| **Priority training needs of healthcare personnel** |  |  |
| National pharmacovigilance regulation | 148 | 69.2 |
| International pharmacovigilance regulation | 109 | 50.9 |
| Organization of the pharmacovigilance system in Cameroon | 144 | 67.3 |
| Case detection | 144 | 67.3 |
| Case notification | 130 | 60.7 |
| Data analysis | 104 | 48.6 |
| Causality assessment | 98 | 45.8 |
| Good Clinical Practices in Pharmacovigilance | 131 | 61.2 |
| Other | 16 | 7.5 |
| **Most accessible training sources for healthcare personnel** |  |  |
| Online courses | 94 | 43.9 |
| Workshop | 154 | 72.0 |
| Internship | 79 | 36.9 |
| Other | 9 | 4.2 |
| **Challenges encountered by healthcare personnel during pharmacovigilance activities** |  |  |
| Lack of training on pharmacovigilance implementation procedures | 125 | 58.4 |
| Limited awareness of the pharmacovigilance system | 86 | 40.1 |
| Lack of pharmacovigilance guidelines | 126 | 58.9 |
| Lack of adverse event reporting forms | 112 | 52.3 |
| Insufficient resources to conduct pharmacovigilance investigations | 82 | 38.3 |
| Lack of motivation to perform pharmacovigilance activities | 24 | 11.2 |
| Lack of supervision | 29 | 13.5 |
| Lack of feedback | 38 | 17.8 |
| Uncertainty about where to report adverse events | 24 | 11.2 |
| Other | 36 | 16.8 |

## Discussion

This study identified important gaps in the organization and implementation of pharmacovigilance activities in participating health facilities, as well as substantial training needs among healthcare personnel. Only 40.0% of health facilities had an established pharmacovigilance unit, and the availability of pharmacovigilance resources and tools was limited. The main challenges reported by health facilities were insufficient budget allocation, limited access to pharmacovigilance training, lack of pharmacovigilance guidelines and insufficient qualified human resources (80.0% each). Among healthcare personnel, previous exposure to pharmacovigilance training was limited, with only 37.9% reporting prior training, while all participants expressed a need for additional training. The main areas identified for further training included national pharmacovigilance regulations, the organization of the pharmacovigilance system in Cameroon, and adverse event detection.

Health facilities play a central role in pharmacovigilance because they prescribe, dispense, and administer medicines and are often the first point of contact for patients experiencing adverse drug reactions [15]. Consequently, they should have the organizational capacity to detect, document, investigate, and report suspected adverse drug reactions in accordance with national and international pharmacovigilance guidelines. These guidelines recommend establishing dedicated pharmacovigilance structures to coordinate surveillance activities and strengthen medicine safety within health facilities [11, 16]. Our findings showed that only four of the ten participating health facilities had an established pharmacovigilance unit, indicating limited institutional capacity to support pharmacovigilance activities. This finding is consistent with a study conducted in Morocco, where fewer than half of hospitals had implemented pharmacovigilance activities [17]. Furthermore, the absence of pharmacovigilance units in some referral hospitals is of particular concern, given their role in managing patients with complex conditions and administering high-risk medicines. These findings suggest important weaknesses in the institutional implementation of pharmacovigilance activities and raise concerns about the capacity of health facilities to effectively monitor medicine safety. Strengthening institutional support for pharmacovigilance, including the establishment and operationalization of pharmacovigilance units, particularly in referral and tertiary-level hospitals, may improve the detection, reporting, and management of adverse drug reactions.

Adequate infrastructure and resources, including dedicated office space, trained personnel, computers, and appropriate operational tools, are essential for the effective implementation of pharmacovigilance activities such as adverse event detection, reporting, investigation, supervision, and staff training. The present study revealed substantial deficiencies in the availability of these resources across participating health facilities, with half of the facilities reporting no pharmacovigilance resources and only one facility having data management tools, including a computer and data archiving system. Such resource constraints are likely to limit the capacity of health facilities to implement routine pharmacovigilance activities and maintain functional surveillance systems. Although the underlying reasons for these deficiencies were not investigated, they underscore the need to strengthen institutional capacity for pharmacovigilance. Ensuring the availability of essential infrastructure, operational resources, and dedicated funding, particularly in referral and regional hospitals, could substantially improve the implementation and sustainability of pharmacovigilance activities.

Health facilities represent the primary setting for identifying and managing suspected adverse drug reactions and therefore play a pivotal role in pharmacovigilance. They are expected to perform core pharmacovigilance activities, including adverse event detection, reporting, investigation, causality assessment, and data analysis. However, this study revealed limited implementation of these activities, with 30% of health facilities conducting neither pharmacovigilance detection nor notification activities. Although comparable studies evaluating the implementation of individual pharmacovigilance activities at the health facility level are scarce, these findings are consistent with broader evidence from low- and middle-income countries showing that limited institutional capacity, inadequate training, and weak reporting systems remain major barriers to effective pharmacovigilance [18, 19]. Strengthening the implementation of core pharmacovigilance activities across health facilities is essential to improve the early detection, reporting, investigation, and assessment of adverse drug reactions. This is especially important in referral and tertiary-level hospitals, where the use of high-risk medicines, such as anticancer agents, may increase the likelihood of serious adverse drug reactions and where specialized expertise can support pharmacovigilance activities at lower levels of the health system.

Healthcare personnel are the cornerstone of patient care and play a central role in pharmacovigilance within health facilities. As the first point of contact for patients experiencing suspected adverse drug reactions, they are expected to possess a sound understanding of the fundamental principles of pharmacovigilance. The present study identified important gaps in pharmacovigilance knowledge among healthcare personnel. More than half of the participants were unable to correctly define an adverse event. Although most participants understood the importance of reporting adverse events and were aware of the recommended reporting timeframe and knowledge of reporting procedures remained suboptimal. These gaps may result in missed or delayed reporting of suspected adverse drug reactions, thereby limiting the effectiveness of pharmacovigilance systems [16, 20]. Similar deficiencies in pharmacovigilance knowledge among healthcare personnel have been reported in other settings [21, 22]. The limited previous exposure to pharmacovigilance training observed in this study, together with the limited availability of pharmacovigilance guidelines in participating health facilities, may partly explain these findings. Strengthening both pre-service and in-service pharmacovigilance training, alongside the provision of practical guidance and standard reporting tools, could improve the quality and completeness of adverse event reporting. Integrating pharmacovigilance more comprehensively into the curricula of medical, pharmacy, nursing, and other health sciences training programs may also help ensure that future healthcare professionals enter clinical practice with the competencies required to detect, assess, and report adverse drug reactions effectively.

Only 37.9% of healthcare personnel reported having previously received pharmacovigilance training, indicating limited exposure to formal training in medicine safety surveillance. Among those who had received training, the most frequently covered topics were adverse event detection, adverse event reporting, and Good Clinical Practice (GCP) in pharmacovigilance, whereas key areas such as causality assessment, data analysis, and the organization of the national pharmacovigilance system were addressed less frequently. Similar levels of pharmacovigilance training have been reported among healthcare professionals in other African countries. Limited training has been identified as an important contributor to the underreporting of adverse drug reactions by reducing healthcare professionals’ ability to recognize and report suspected adverse events, while other barriers described in the literature include limited awareness of reporting requirements, inadequate access to reporting forms, fear of blame or litigation, and uncertainty regarding causality [23]. These findings are of particular concern given that adverse drug reactions remain one of the major causes of morbidity, hospitalization, and avoidable healthcare burden worldwide.

The organizational and resource gaps identified at the health facility level were consistent with the training needs and implementation challenges reported by healthcare personnel, suggesting a shared recognition of the key barriers to effective pharmacovigilance. This concordance provides an opportunity to develop targeted interventions that address both institutional and workforce-related constraints. In addition to the need for training, respondents highlighted the importance of strengthening staff motivation, improving adverse event reporting, and increasing stakeholder engagement in pharmacovigilance activities. Similar priorities have been reported among healthcare professionals in other settings [22, 24]. These findings highlight the importance of adopting comprehensive strategies that address organizational capacity, human resources, and continuous professional development to strengthen pharmacovigilance systems and improve medicine safety surveillance.

### Study limitations

This study has several limitations. First, it was conducted exclusively in referral and regional hospitals, which may limit the generalizability of the findings to primary-level health facilities and other healthcare settings. Second, although most eligible healthcare personnel participated, not all targeted health facilities and personnel consented to participate, raising the possibility of selection bias. Third, the use of a questionnaire-based survey may have introduced information bias, including recall and social desirability bias, as responses were self-reported rather than independently verified. Despite these limitations, this study provides valuable insights into the organization of pharmacovigilance systems, the knowledge of healthcare personnel, and the training needs and implementation challenges within referral health facilities.

These findings provide an evidence base to inform strategies aimed at strengthening pharmacovigilance capacity and improving medicine safety surveillance in similar settings.

## Conclusion

Pharmacovigilance implementation in referral and regional health facilities in Cameroon remains limited, with important gaps in organizational structures, the availability of essential resources and tools, healthcare personnel’s knowledge, and training. Although pharmacovigilance activities were reported in most participating facilities, only a minority had the resources and tools required to conduct these activities effectively. Furthermore, fewer than half of healthcare personnel had previously received pharmacovigilance training, and substantial knowledge gaps were identified. Strengthening pharmacovigilance systems therefore requires a comprehensive approach that combines enhanced facility-level capacity and resources, improved availability and dissemination of standardized guidelines and reporting tools, regular and targeted training of healthcare personnel, and strengthened supervision, data analysis, and feedback mechanisms. These measures are essential to improve the detection and reporting of adverse events, promote the effective use of safety information, and ultimately enhance medicine safety and patient care in Cameroon.

## Data Availability

Data presented or used for the analysis in the present manuscript are available as ‘Supplementary material 1’.

## List of Legends

Supplementary material 1: Database.

## References

[1] Curtin F, Schulz P. Assessing the benefit: risk ratio of a drug--randomized and naturalistic evidence. Dialogues Clin Neurosci. 2011;13(2):183–90.

[2] Muñoz LM. Introduction to Post-marketing Drug Safety Surveillance: Pharmacovigilance in FDA/CDER 2016 [Available from: https://www.fda.gov/files/about%20fda/published/Introduction-to-Post-Marketing-Drug-Safety-Surveillance-%28PDF---1.46MB%29.pdf.

[3] Suvarna V. Phase IV of Drug Development. Perspect Clin Res. 2010;1(2):57–60.

[4] WHO. The IMPORTANCE of PHARMACOVIGILANCE 2002 [Available from: https://iris.who.int/server/api/core/bitstreams/002b78d5-4dfc-433c-a518-f92ebdee8706/content.

[5] WHO. Tools and Innovations in Pharmacovigilance [Available from: https://www.who.int/teams/regulation-prequalification/regulation-and-safety/pharmacovigilance/guidance/operations/tools-innovations.

[6] WHO. How to Set Up a Pharmacovigilance (PV) Centre [Available from: https://www.who.int/teams/regulation-prequalification/regulation-and-safety/pharmacovigilance/guidance/operations/pv-center.

[7] WHO. More Global commitment needed to monitor Safety and Quality of medicines [Available from: https://www.who.int/news/item/14-10-2002-more-global-commitment-needed-to-monitor-safety-and-quality-of-medicines.

[8] WHO. Patient safety 2023 [Available from: https://www.who.int/news-room/fact-sheets/detail/patient-safety.

[9] Miguel A, Azevedo LF, Araújo M, Pereira AC. Frequency of adverse drug reactions in hospitalized patients: a systematic review and meta-analysis. Pharmacoepidemiol Drug Saf. 2012;21(11):1139–54.

[10] Nyame L, Hu Y, Xue H, Fiagbey EDK, Li X, Tian Y, et al. Variation of adverse drug events in different settings in Africa: a systematic review. Eur J Med Res. 2024;29(1):333.

[11] Monitoring WCCfID. Safety Monitoring of Medicinal Products: Guidelines for Setting Up and Running a Pharmacovigilance Centre 2000 [Available from: https://books.google.cm/books/about/Safety_Monitoring_of_Medicinal_Products.html?id=k_vnjwEACAAJ&redir_esc=y.

[12] Presidency of the Republic of Cameroon. DECREE N°2013/093 of April 03, 2013 on the organization of the Ministry of Public Health 2013 [Available from: http://cdnss.minsante.cm/?q=fr/content/decret-n-2013093-du-03-avril-2013-portant-organisation-du-ministère-de-la-santé-publique.

[13] Cameroun MSD. Lignes directrices en matière de pharmacovigilance au Cameroun 2018 [Available from: https://dpml.cm/index.php/fr/publications/guide-de-bonnes-pratiques/pharmacovigilance/396-lignes-directrices-en-matiere-de-pharmacovigilance-au-cameroun.

[14] MINSANTE. Annuaire: Formations Sanitaires Publiques Du Cameroun 2022 [Available from: https://fr.scribd.com/document/768959569/001-069.

[15] Najafi S. Importance of Pharmacovigilance and the Role of Healthcare Professionals. Journal of Pharmacovigilance 2018;06(01).

[16] MINSANTE. LIGNES DIRECTRICES EN MATIERE DE PHARMACOVIGILANCE AU CAMEROUN 2018 [Available from: https://dpml.cm/images/Publications/GuideBonnePratique/LignesDirectricesPharmacovigilance_DPML_Cameroun.pdf.

[17] Hamzaoui H, Shaum A, Cherkaoui I, Moussa LA, Sefiani H, Talibi I, et al. Assessment of Pharmacovigilance Across University Hospitals in Morocco. Drug Saf. 2025;48(5):527–39.

[18] Avong YK, Jatau B, Gurumnaan R, Danat N, Okuma J, Usman I, et al. Addressing the under-reporting of adverse drug reactions in public health programs controlling HIV/AIDS, Tuberculosis and Malaria: A prospective cohort study. PLoS One. 2018;13(8):e0200810.

[19] Varallo FR, Guimarães Sde O, Abjaude SA, Mastroianni Pde C. [Causes for the underreporting of adverse drug events by health professionals: a systematic review]. Rev Esc Enferm USP. 2014;48(4):739–47.

[20] WHO. WHO pharmacovigilance indicators: a practical manual for the assessment of pharmacovigilance systems 2015 [Available from: https://www.who.int/publications/i/item/9789241508254.

[21] Adisa R, Omitogun TI. Awareness, knowledge, attitude and practice of adverse drug reaction reporting among health workers and patients in selected primary healthcare centres in Ibadan, southwestern Nigeria. BMC Health Serv Res. 2019;19(1):926.

[22] Olsson S, Pal SN, Stergachis A, Couper M. Pharmacovigilance activities in 55 low- and middle-income countries: a questionnaire-based analysis. Drug Saf. 2010;33(8):689–703.

[23] Isah AO, Pal SN, Olsson S, Dodoo A, Bencheikh RS. Specific features of medicines safety and pharmacovigilance in Africa. Ther Adv Drug Saf. 2012;3(1):25–34.

[24] Wilbur K. Pharmacovigilance in Qatar: a survey of pharmacists. East Mediterr Health J. 2013;19(11):930–5.

